# Biases in the Willingness to Download a mHealth App: A Discrete Choices Experimental Study Among Nigerian Healthcare App Subscribers

**DOI:** 10.1101/2024.01.08.24300983

**Authors:** Ignatius Nwoyibe Ogbaga

## Abstract

**Background:** Although there has been an increase in the availability of mobile health (mHealth) tools globally and their potential benefits for both healthcare providers and patients, the adoption of mHealth is still relatively low. Additionally, only a limited number of studies have investigated the intention of individuals to download mHealth apps.

**Objective:** We conducted a study to explore people’s inclination towards using a health app.

**Methods:** We conducted the study in Nigeria using a discrete choice experiment. The study had a sample size of 2800 participants who were presented with two different attributes and levels. These attributes were price ($20 = N 17932.80 [at a currency exchange rate of $1= N896.64], and free subscription option) and data protection (with options of data protection vs no data protection). The participants were randomly assigned to the different attribute and level options. For the analysis, we used the conditional logistic model.

**Results:** According to the results of the study, the likelihood of downloading a mHealth app is significantly higher when the app is offered for free. The study also found that users tend to ignore data protection specifications, and instead prioritize free subscription offers while showing reluctance towards apps that come with a price tag.

**Conclusions:** The use of mobile health (mHealth) tools has a high potential in reducing healthcare costs and enhancing the efficacy of traditional health interventions and therapies. The major driving forces behind the increasing adoption of mHealth apps in the future are cost reduction and the establishment of sound business models. It is crucial to establish reliable standards for mHealth apps, which can include information about pricing and legislation regarding data protection, to ensure that potential consumers can make informed decisions.

## Introduction

### Background

Mobile health (mHealth) aims to enhance the quality of healthcare, increase its accessibility, and reduce costs associated with healthcare by leveraging mHealth apps in remote healthcare delivery (Silver *et al*., 2015), mHealth apps are computer programs designed to run on mobile devices like smartphones and tablets to support health and health-related behaviours (Smit and Bol, 2019). Studies have shown that mHealth could improve health and well-being globally by reducing healthcare costs, improving healthcare quality, and promoting behaviour change to help prevent diseases, including chronic diseases. However, despite the potential benefits of mHealth apps for improving health and lifestyle, their actual adoption and long-term use are rather low, lagging behind their potential (Tomlinson *et a*l., 2013). Therefore, there is insufficient evidence to inform the implementation and scale-up of mHealth apps, despite their promising effects on health monitoring and improvement (Kumar *et al*., 2013). To address this issue, it is crucial to understand individual preferences that predict the willingness to download mHealth apps (Anastasiadou *et al*., 2018).

With the rapid development of mobile technologies, delivering behaviour change interventions through mHealth apps has become increasingly popular. However, health communication researchers need to further improve their understanding of how to advance current theories so that we can make the most of the ubiquity, adaptability, and affordability of these apps in changing health behaviours (Ng JYY *et al*., 2012). To begin this effort, it is important to understand how various attributes of mHealth apps impact people’s willingness to download them. Since there is limited knowledge about the cognitive factors that drive the adoption of such apps, it is necessary to investigate which specific attributes are favoured by users in the adoption process (Folkvord *et al*., 2022). Intrinsic motivation is a strong predictor of mHealth app adoption and usage (Ng JYY *et al*., 2012). It will be difficult to establish the effectiveness of mHealth apps and understand individuals’ use without comprehending cognitive motivators for mHealth app adoption.

### Factors Influencing the Adoption of Mobile Health Apps

The increase in the usage of the mHealth apps market has continued to pose existential threats to the management of health-related data in years to come. Currently, there is a paradigm shift from mainframe systems located in the facilities of healthcare providers to apps on mobile phones and data stored in shared cloud services (He *et al*., 2014). Crucially, the adoption rates of health applications are influenced by factors such as cost and privacy considerations. Notably, individuals’ perceptions regarding their personal information’s confidentiality have a direct correlation with their use of mobile health (mHealth) apps. Those who express greater apprehension about the potential exploitation of their data are less inclined to utilize certain mHealth apps (Bol *et al*., 2018). Additionally, free-of-charge mHealth apps tend to be more attractive to consumers; nevertheless, they frequently incorporate alternative revenue streams like advertising or trading personal information (including sensitive health-related data) (Sax *et al*., 2018)

Moreover, healthcare providers are regarded as the guardians of healthcare delivery (Bernstein *et al*., 2007). Concurrently, with the increasing usage of online platforms, reviews and recommendations have emerged as a crucial source of information that aids consumers in making informed purchase decisions (Zhang *et al*., 2014). Zhang has also formulated a heuristic-systematic model to scrutinize the impact of online reviews and recommendations on consumption behaviour. They demonstrated that both the informativeness and persuasiveness of reviews and recommendations are pivotal attributes that argument quality when deciding whether or not to consume a product. People rely on heuristic information processing while selecting products, which implies that they consider only one or a few informational cues before forming their judgment (Tam and Ho, 2005). In a recent study, COVID-19-tracking apps and healthcare apps from various countries were evaluated using qualitative content analysis according to their reward mechanisms based on the Mobile Application Rating Scale approach for assessment (Peschke 2022). The evaluation included an examination of different rewards for voluntary participation. The Mobile Application Rating Scale approach encompasses engagement, functionality aesthetics, and information quality. It is noteworthy that motivational strategies such as gamification techniques and tools for individual knowledge exchange lower the inhibition threshold associated with downloading and utilizing healthcare apps along with COVID-19-tracking apps.

Drawing from the principle of least effort, individuals generally prefer to minimize cognitive exertion and only expend significant effort when necessary (Bohner *et al*., 1995). For instance, if a healthcare professional, such as a physician, recommends a mHealth application, people are more likely to perceive this guidance as credible when deciding whether or not to adopt and utilize it (Chen and Chaiken, 1999). There are at least two reasons why a doctor’s endorsement of a mHealth app can serve as a powerful motivator for patients seeking to incorporate digital health technologies into their lives. Doctors are recognized experts in their respective fields and therefore wield greater influence than non-experts - particularly since they possess intimate knowledge of their patients’ interests (Leigh and Ashall-Payne, 2019).

Conclusively, the manufacturer of a mHealth application can act as a heuristic in the adoption process by consumers. There is reason to believe that this could be the case, considering the pharmaceutical industry has faced challenges with public perception over recent decades. Pharmaceutical companies must navigate a delicate balance between pursuing optimal healthcare and striving for profit (Bauchner and Fontanaros, 2013).

### The Study

We undertook a large-scale Discrete Choice Experiment (DCE) in Nigeria to compare the price of free subscription apps versus paid subscriptions (at an exchange rate of $20 = N 17932.80 and $1= N896.64) along with data protection options to determine the likelihood of people adopting mHealth applications. This study delved into the diverse cultural backgrounds of participants and their local healthcare facilities.

Infrastructure-wise, Nigeria has a National Health Insurance Scheme (NHIS), which is mandated for private insurance on a national level where the government regulates and subsidizes insurance for subscribers. However, state and local governments are not statutory members of this scheme as they have their separate insurance programs. Given these differences in healthcare systems, our experiment aimed to ascertain user preferences comprehensively.

Our objective was to provide an all-encompassing evaluation of individual preferences towards mHealth application adoption. To our knowledge, no prior research has endeavoured to differentiate between preferences for mHealth app adoption using DCEs in Nigeria. Therefore, insights into attributes that could influence individuals’ likelihood to adopt a mHealth app would be highly beneficial in informing product development and pathway redesign for future technological advancements within the realm of mobile health services.

## Methods

### Design

In this research, we employed a discrete choice experiment (DCE) through an online questionnaire. In contrast to other preference elicitation techniques, a DCE is capable of quantifying the relative importance of diverse attributes that define a novel or preexisting product or service. This method identifies which attributes people prioritize or accept and which they may be willing to exchange to optimize their utilization (Lancsar, 2008). A DCE necessitates participants to select between competing scenarios, such as costs described in terms of a specific attribute (e.g., price, ease of use, ease of access, etc). By offering participants with distinct attributes and levels within those attributes and subsequently requesting them to make decisions regarding their preferred option over another, researchers can ascertain the most favoured combination of options in participant decision-making. The relevance of DCE studies lies in their ability to enable direct cognitive assessment of relative preferences for various existing and hypothetical new service configurations or treatment approaches. Moreover, by utilizing a DCE approach, it becomes possible to evaluate directly the complexity involved in human decision-making by presenting small variations in the options presented to participants.

In this study, the online questionnaire utilized a main effects design that incorporated all attributes examined. The main effects design encompasses all feasible levels and ensures an equal occurrence of each level pair. A full factorial design was avoided in favour of a fractional factorial design since the former would have entailed too many possible alternatives, rendering it impractical for individuals to complete or for a blocked questionnaire format to be managed effectively.

The utilization of an electronic survey provided data on completion time, which facilitated the internal validity checks and provided a precise account of the duration taken to finish the questionnaires. Two sets of cognitive evaluations (n=48) were conducted to assess respondents’ understanding levels when making decisions. These preliminary tests validated that utilizing the questionnaire as a basis for the study was satisfactory and comprehensible for participants after some minor revisions in the task explanation. The 4 characteristics (price and data confidentiality) and levels picked for integration in the DCE are displayed in Table 1.

| **CODIFICATION** | **DESCRIPTION** |
| --- | --- |
| **Subscription Cost** |  |
| P1 | Paid (\$20 = N 17932.80 at \$1= N896.64) |
| P2 | For free |
| **Data protection** |  |
| D1 | Data protection |
| D2 | No information |

### Procedure

All the survey respondents were duly intimated on the research objectives and procedures. It was only those who consented to partake in the study were given access to the online questionnaire platform.

Initially, the participants were requested to furnish their socio-demographic details, such as age, gender, education and employment status. Following this, an introduction was provided on the attributes and levels of the DCE questionnaire. The questionnaire design involved selecting a generic pairwise choice along with an opt-out question (“Not interested in the app download”). The participants were presented with numerous sets of choices where they could select from three responses: “Option A,” “Option B,” or “I would not download the app.” A sample choice set is illustrated in Table 2

**Table 2.** A prototype of the discrete decision-making exercise.

| Options | Item | Condition of use |
| --- | --- | --- |
| Option A | The price for the app is \$20 " (a currency exchange rate of \$1= N896.64 is applicable | No information regarding the utilization of the data supplied into the application |
| Option B | "The app is for free" | The app's handling of user-entered data and its subsequent storage remains undisclosed to the users |
| Not downloaded | "I would not download the app" |  |

### Participants

The data used for this study was collected from Nigerians living in Nigeria. A total of (n=1600) participants participated in the online survey. The sample was chosen through a proportionate stratified sampling method, considering gender and age.

### Ethical Considerations

This research received ethical clearance from the Ebonyi State University, Abakaliki ethical committee. All participants provided written and informed consent voluntarily without receiving compensation. By signing the informed consent form, individuals were guaranteed that their data would be kept confidential and that they could withdraw from the study at any time.

### Statistical Analyses

We analyzed the choice experiment data using a conditional logit model that was estimated through the use of Clogit R software, which was developed by the R Foundation for Statistical Computing. The estimation process required that we prepare a data structure containing one row for each alternative presented to decision-makers in every choice scenario. For each alternative, we utilized a dependent variable coded as a dummy variable with a value of 1 if it was selected by respondents. Given that participants were exposed to 18 unique scenarios and three alternatives (two apps and an opt-out option) within each scenario, our dataset consisted of 43,200 rows - specifically (1600x18x3: where there are 1600 participants x 18 choices available x 3 alternatives).

*Kij* = β1 alternative specific constant + β2 price_200 + β3 data protection

Each regressor denotes an indicator variable signifying the presence of a particular feature in the application. The alternative specific constant signifies which option among each choice task serves as the opt-out option. Consequently, its coefficient estimates whether individuals were inclined towards not downloading the app. We included the “not to download” option to evaluate participants’ preferences regarding app downloads without imposing it on them to make that choice.

## Results

Table 3 displays the participant characteristics. The descriptive data indicates that the majority of participants belong to a young student population with higher levels of education, lower financial status, lower scores on health consciousness, and a high orientation towards health information.

**Table 3.** Descriptive information about the participants per country.

| Characteristics | Participants (n=1600) |
| --- | --- |
| Gender: Male, n (%) | 900 (56.25%) |
| Gender: women, n (%) | 700 (43.75%) |
| Age (years), mean (SD)<sup>a</sup> | 61.60 (33.32) |
| **Educational level, n (%)<sup>a</sup>** |  |
| Primary education | 35(2.19%) |
| High school diploma | 260(16.25%) |
| University undergraduate | 500(31.25%) |
| Graduate | 305(19.06%) |
| Postgraduate degree | 200(12.50%) |
| Non-students | 300(18.75%) |
| **Employment status, n (%)<sup>a</sup>** |  |
| Employed | 400(25%) |
| Unemployed | 200(12.50%) |
| Student | 600(37.50%) |
| Retired | 50(3.13%) |
| Other Reasons | 350(21.88%) |
| **Difficulty paying a subscription fee, n (%)<sup>a</sup>** |  |
| Always | 350(21.88%) |
| periodically | 650(40.63%) |
| Rarely | 200(12.50%) |
| No Response | 400(25%) |
| **Health app use, n (%)** |  |
| Non Use | 320(20%) |
| Once | 400(25%) |
| Twice | 325(25%) |
| 3 times | 85(5.31%) |
| 4 times | 250(15.63%) |
| 5 times | 140(8.75%) |
| 6 or more times | 80(5%) |
| **Health in general, n (%)<sup>a</sup>** |  |
| Very bad | 1(0.1%) |
| Bad | 42(5.20%) |
| Neutral | 258(32.20%) |
| Good | 922(115.20%) |
| Very good | 376(47%) |

<sup>a</sup> $P < .001$ .

Table 4 displays the outcomes of the conditional logistic model examination of the gathered data, where results are presented in odds ratios (OR). Thus, values exceeding 1 indicate an increase in the probability of downloading this app compared to the reference group.

**Table 4.** Conditional logistic model for the discrete choice experiment outcomes among participants.

| Variable | Dependent variable: Choice |
| --- | --- |
| **Price \$20<sup>a</sup>** | |
| OR<sup>b</sup> (95% CI) | 0.498<sup>c</sup> (0.468-0.530) |
| SE | 0.064 |
| **Data protection** |  |
| OR (95% CI) | 0.25<sup>c</sup> (0.232-0.270) |
| SE | 0.078 |
| **Alternative specific constant** |  |
| OR (95% CI) | 0.675<sup>c</sup> (0.627-0.728) |
| SE | 0.038 |
| Observations | 43,200 |
| $R^2$ | 0.113 |
| Maximum possible $R^2$ | 0.697 |
| Log-likelihood | -23,165.480 |
| Wald test | 4127.660<sup>c</sup> |
| Likelihood ratio test | 5197.001<sup>c</sup> |
| Score (log-rank) test | 4847.258<sup>c</sup> |

<sup>a</sup>A currency exchange rate of \$1=N896.64 is applicable.
<sup>b</sup>OR: odds ratio.
<sup>c</sup> $P < .01$ .

To begin with, participants were more inclined to download free applications. This is consistent with the findings of Folkvord where Dutch users exhibit less sensitivity towards priced apps (Folkvord et al.,2023).

Subsequently, the respondents expressed a favourable appreciation for applications that guarantee the protection of confidential information.

Lastly, our findings suggest that respondents are more inclined towards abstaining from app downloads when there is no discrete data protection information attached to the app.

## Discussion

### Principal Findings

This research was conducted through a discrete choice experiment (DCE) in which we systematically varied the price and level of data protection, ultimately examining preferences for downloading a mHealth application by users. Our objective was to perform a thorough evaluation of individuals’ inclinations towards adopting such an app. DCE methodology necessitates participants selecting between competing scenarios and comparing them with alternative options, providing greater insight into people’s preferences.

The study revealed that the cost and safeguarding of personal information were pivotal determinants in shaping individuals’ inclination to acquire the mHealth application. Download rates increased substantially when the app was offered for free, aligning with (Folkvold et al., 2023) research findings in Spain, Germany, and the Netherlands; where participants exhibited a greater likelihood of downloading health apps that were free of charge. Despite their nominal value, these factors had a significant impact on download probability. Furthermore, the majority of the participants expressed more willingness to download an app if its data privacy was protected by law than otherwise.

Generally, individuals tend to rely on heuristic information processing when making decisions about product consumption. This involves applying the least amount of effort by considering only one or a few informational cues to form a judgment (Tam and Ho, 2005). This research demonstrates that price and data protection are particularly significant factors for determining whether an individual is willing to download a health app or not.

Drawing from the health belief model (Ahadzadeh et al., 2015) and technology acceptance model (Yuen et al., 2020), people are unlikely to engage in health or preventative measures unless they have a strong conviction to utilize mHealth applications, believing them to be both important and beneficial to them. Factors such as cost and privacy serve as important determinants that either facilitate or impede the adoption of a mHealth app (Champion and Skinner, 2008). For this reason, verified guidelines and dependable efficacy are imperative.

The major driving forces for adopting mHealth in the future are improving patient safety (data protection), reducing costs, and creating sound business models. To ensure that users can confidently use mobile apps that are of high quality, take care of data protection issues and reflect their value, it is essential to create standards for mobile apps. Governments, funders, and industry associations should create and implement such standards.

It would be helpful if healthcare professionals, institutes, and organizations established guidelines to assist developers in creating high-quality and valuable mobile health (mHealth) applications. These guidelines could cover safety, accuracy, and security aspects. Based on these factors, clinicians might recommend specific apps and utilize the clinical data that these apps provide to make medical decisions

### Strengths and Limitations

It is important to note that there has been no research on the factors that influence the adoption of mHealth apps in Nigeria. We believe that, identifying these factors could provide valuable insights for product development and redesigning of future mHealth technologies. We also analyzed some key factors that affect the adoption of mHealth apps and tested various attributes that are important in predicting and explaining the adoption of these apps.

The study was not without its shortcomings. primarily because the study was done online, it is impossible to guarantee the internal validity of the exact experiment because of the uncertainty around the seriousness of the participants’ participation. Nevertheless, employing an online questionnaire to evaluate various characteristics can be regarded as a valid and trustworthy method because the experiment focused on factors that predict the adoption of a mHealth app, which can be considered an online behaviour.

## Conclusions

The global population of smartphone users for health-related reasons has grown significantly during the past 20 years (Gagnon et al., 2016). Adopting a mHealth app is a must for effectiveness, but research on the motivating factors driving mHealth adoption is still lacking (Böhm et al., 2019). The findings from this research can be applied by a variety of stakeholders who are working on creating mHealth technologies. The reasons that will propel the use of mHealth in the future are enhancing patient safety (data protection), price reduction, and developing strong business models (Chan et al., 2017). For mobile apps, it is therefore necessary to establish reliable standards and norms; in particular, data protection laws.

## Acknowledgements

We sincerely acknowledge the authors whose works we cited. This research was not supported by any funder

## Data Availability

The data sets generated during and analyzed during this study are available.on reasonable request to the corresponding author, it will be made available.

## Declaration of generative AI and AI-assisted technologies in the writing process

During the preparation of this work, the author(s) used Grammarly and WPS AI tools to improve the readability of our material. After using this tool/service, the author reviewed and edited the content as needed and took full responsibility for the content of the publication.

